# GPR143, a novel immunohistochemical marker for renal tumors with *FLCN/TSC/MTOR-TFE* alterations

**DOI:** 10.64898/2026.04.06.26350070

**Authors:** Qinyuan Li, Anupama Singh, Rong Hu, Wei Huang, Daniel D. Shapiro, E. Jason Abel, Yang Zong

**Author notes:** To whom correspondence may be addressed: Dr. Yang Zong, Department of Pathology and Laboratory Medicine, University of Wisconsin-Madison, 600 Highland Avenue, Madison, WI 53792, USA.

## Abstract

Although several ancillary tests are available in limited laboratories, diagnosis of microphthalmia (MiT)/TFE family translocation renal cell carcinoma (tRCC) could be challenging due to diverse and overlapping tumor morphology and the lack of reliable biomarkers. GPNMB has been recently identified as a diagnostic marker for various renal neoplasms with *FLCN/TSC/mTOR-TFE* alterations. However, the sensitivity and specificity of GPNMB immunostain are suboptimal and the result interpretation in ambiguous cases could be difficult. To search additional biomarkers that could improve the screening sensitivity and predict genetic aberrations in *FLCN/TSC/mTOR-TFE* pathway in renal tumors, we performed bioinformatic analysis of publicly available cancer databases and found GPR143, a transmembrane protein regulated by MiT transcription factors, was highly expressed in a subset of renal cell carcinomas (RCCs). In two the Cancer Genome Atlas (TCGA) kidney cancer cohorts, RCCs with high levels of *GPR143* expression were enriched for renal neoplasms with *FLCN/TSC/mTOR-TFE* alterations. Similar to GPNMB labeling, GPR143 immunostain was positive in the majority of tRCC cases and renal tumors with *FLCN/TSC/mTOR* alterations, suggesting that GPR143 could function as another surrogate marker for *FLCN*/*TSC/mTOR-TFE* alterations in certain renal tumors. Interestingly, despite the concordant GPR143 and GPNMB immunoreactivity in most renal neoplasms with *FLCN/TSC/mTOR-TFE* alterations, diffuse GPR143 immunostain was observed in some cases with negative or focal GPNMB labeling. Taken together, our results indicate GPR143 could serve as a useful adjunct marker to improve the sensitivity for screening renal tumors with *FLCN/TSC/mTOR-TFE* alterations.

## Introduction

GPNMB (Glycoprotein nonmetastatic melanoma protein B), a transmembrane protein regulated by the microphthalmia (MiT)/TFE family of transcription factors, has been recently discovered as a diagnostic biomarker for various renal neoplasms with *FLCN/TSC/mTOR-TFE* alterations.^1,2^ High expression of GPNMB was found in MiT/TFE family translocation renal cell carcinoma (tRCC),^1-3^ renal tumors associated with Birt–Hogg–Dubé (BHD) syndrome or somatic *FLCN* mutations,^4-6^ and many renal tumors associated with *TSC/mTOR* signal pathway aberrations, including renal cell carcinoma (RCC) with fibromyomatous stroma,^7-9^ eosinophilic solid and cystic renal cell carcinoma (ESC RCC), low grade oncocytic tumor (LOT) and eosinophilic vacuolated tumor (EVT)^1,2,6^ as well as angiomyolipoma.^1,2^ However, approximately 13% - 30% of tRCC cases are negative for diffuse GPNMB staining.^1,2^ Furthermore, a large fraction of chromophobe renal cell carcinoma (chRCC) show diffuse or partial positive immunoreactivity with GPNMB antibody.^1,2,6^ Several other renal epithelial tumors including oncocytoma, clear cell renal cell carcinoma (ccRCC) and papillary renal cell carcinoma (pRCC) could rarely exhibit diffuse or focal staining for GPNMB.^2,5,6^

Given that many renal neoplasms with *FLCN/TSC/mTOR-TFE* alterations also express several downstream targets of the MiT/TFE family of transcription factors, such as Melan-A, HMB45, cathepsin K and TRIM63,^10-13^ we postulated that immunohistochemistry (IHC) analysis with antibodies against additional MiT/TFE downstream targets could provide useful adjunct tests to existing markers and may improve the sensitivity and specificity for the diagnosis of renal tumors with *FLCN/TSC/mTOR-TFE* alterations.

GPR143 protein is a unique G protein-coupled transmembrane receptor protein involved in the pathogenesis of ocular albinism.^14^ It has been identified as a downstream target of MiT transcription factors, which is mainly localized in intracellular organelles, such as late endosomes and melanosomes, in pigment cells.^15-17^ In addition to its important role in melanosome biogenesis and maturation, it has recently reported that GPR143 is also implicated in exosome biogenesis and promotes cell mobility and cancer metastasis.^18^ In a genetically engineered mouse model of *TFE3*-rearranged RCC, both GPNMB and GPR143 were significantly overexpressed in mouse kidneys.^19^ In this study, we found that in The Cancer Genome Atlas (TCGA) kidney cancer cohorts, human renal cell carcinomas (RCCs) with high levels of GPR143 expression were enriched for renal neoplasms with *FLCN/TSC/mTOR-TFE* alterations. Similar to GPNMB, IHC analysis with an antibody against GPR143 was positive in the majority of tRCC cases and renal tumors with *FLCN/TSC/mTOR* alterations, suggesting that GPR143 could function as another surrogate marker for *FLCN* mutations and *TSC/mTOR-TFE* signal pathway aberrations in certain renal tumors.

## Materials and methods

### Bioinformatics Tool and Data Analysis

To examine the expression of GPR143 in RCCs, we performed *in silico* data analysis using the following open-access cancer research databases and interactive web portals: 1. The Human Protein Atlas (HPA) database (http://www.proteinatlas.org), which is the most comprehensive database for evaluating the distribution of all the human proteins in normal cells, tissues and organs as well as major cancer types.^20^ The mRNA data of GPR143 expression from 21 cancer types and 3 major RCC subtypes were obtained through the HPA database. 2. cBioPortal database (https://www.cbioportal.org/) is a web-based public platform for interactively exploring multidimensional cancer genomics datasets.^21^ Specific gene fusions, gene mutations, amplifications or other genetic changes of the *FLCN/TSC/mTOR-TFE* pathway in RCCs were identified by manual inquiry in TCGA datasets (TCGA-KIRC, TCGA-KIRP and TCGA-KICH cohorts) via cBioPortal.

### Case Selection

We retrospectively reviewed the electronic medical record from our surgical pathology laboratory registry in the University of Wisconsin Hospital and Clinics and identified 23 renal tumors closely associated with *FLCN/TSC/mTOR-TFE* alterations from 22 patients between 2005 through 2025. The cohort included seven tRCC cases (six *TFE3*-rearranged RCC either diagnosed by histology with unequivocal TFE3 IHC test (n=2) or confirmed by *TFE3* fluorescence in-situ hybridization (FISH) analysis (n=4), and one case of FISH confirmed *TFEB*-rearranged RCC with *MALAT1::TFEB* fusion), four *FLCN* mutated renal tumors (two hybrid oncocytic chromophobe tumors (HOCTs), one unclassified low grade oncocytic renal neoplasm and one ccRCC) from three BHD syndrome patients diagnosed with relevant clinical history and positive germline *FLCN* mutations, one *TSC1*-mutated RCC from a patient with tuberous sclerosis complex (TSC), one ESC RCC case with a germline *TSC1* mutation, two LOTs, two EVTs, and six cases of renal angiomyolipoma (AML) including a patient with clinical TSC syndrome and a patient with a germline *TSC2* deletion (Table 1). All archived slides of these cases were retrieved and reviewed. This study was approved by the Institutional Review Board of School of Medicine and Public Health, University of Wisconsin-Madison.

**Table 1.** Clinicopathological characteristics of 23 renal tumors from 22 patients.

| Case | Sex | Diagnosis | Hereditary cancer syndrome | Germline testing | FISH analysis | TFE3 IHC | GPNUMB IHC | GPR143 IHC |
| --- | --- | --- | --- | --- | --- | --- | --- | --- |
| 1 | F | *TFE3*-rearranged RCC | N/A | N/A | ND | + | DP | DP |
| 2 | F | *TFE3*-rearranged RCC | N/A | N/A | ND | + | DP | DP |
| 3 | M | *TFE3*-rearranged RCC | N/A | N/A | + | + | DP | DP |
| 4 | M | *TFE3*-rearranged RCC | N/A | N/A | + | + | FP | DP |
| 5 | F | *TFE3*-rearranged RCC (Biphasic) | N/A | N/A | + | + | FP | DP |
| 6 | F | *TFE3*-rearranged RCC | N/A | N/A | + | + | Neg | Neg |
| 7 | F | *TFEB*-rearranged RCC | N/A | N/A | + | ND | DP | DP |
| 8 | F | HOCT (n=2) | BHD | *FLCN* c.1286dupA (p.H429Qfs*27) | ND | ND | DP | DP |
| 9 | M | Low grade oncocytic neoplasm | BHD | *FLCN* (c.1285dupC) (p.H429Pfs*27) | ND | ND | DP | DP |
| 10 | M | Clear cell RCC | BHD | *FLCN* c.1285dup (p.His429fs) | ND | Neg | Neg | Neg |
| 11 | F | ESC RCC | N/A | *TSC1* (c.1218C>A) (p.Y406*) | ND | Neg | DP | DP |
| 12 | F | *TSC1*-mutated RCC | TSC | *TSC1* (c.364-2A>G) | ND | Neg | DP | Neg |
| 13 | F | LOT | N/A | Neg | ND | Neg | DP | FP |
| 14 | M | LOT | N/A | ND | ND | Neg | DP | FP |
| 15 | M | EVT | N/A | ND | ND | Neg | DP | DP |
| 16 | F | EVT | N/A | ND | ND | ND | Neg | DP |
| 17 | M | Epithelioid AML | TSC | *TSC2* exons 1-41 deletion | ND | ND | DP | DP |
| 18 | M | AML | TSC | ND | ND | ND | DP | DP |
| 19 | F | AML | N/A | ND | ND | ND | DP | DP |
| 20 | F | Epithelioid AML | N/A | ND | ND | + | DP | FP |
| 21 | F | AML | N/A | ND | ND | Neg | DP | DP |
| 22 | F | Lipid-rich AML | N/A | ND | ND | ND | FP | FP |

F, female; M, Male; N/A, not applicable; ND, not done; DP, diffuse positive; FP, focal positive; Neg, negative.

### Immunohistochemistry analysis

Automated IHC was performed on formalin-fixed, paraffin-embedded whole-tissue sections of 5-μm thickness, using the Ventana Discovery Ultra BioMarker Platform (Ventana Medical Systems, Oro Valley, AZ). In brief, tissue sections were subjected to deparaffinization and heat-induced antigen retrieval with Cell Conditioner 1 (Ventana #950-224) at 95□°C for 64 minutes. Tissue sections were then incubated with primary antibodies against GPR143 (rabbit polyclonal antibody, #PA5-51984, 1:100; Invitrogen, Waltham, MA) or GPNMB (E4D7P rabbit monoclonal antibody, #38313, 1;100; Cell Signaling Technology, Danvers, MA) at 37□°C for 60 minutes. After rinsing with reaction buffer (Ventana #950-300), sections were incubated with Discovery OmniMap anti-rabbit horseradish peroxidase conjugated secondary antibody (Ventana #760-4311) for 16 minutes at 37□°C. Ventana UltraView DAB reagents were applied for detection, visualization and counterstaining. The sections were dehydrated and cover-slipped with permanent media.

IHC results were assessed by two pathologists and categorized as follows: diffuse positive (moderate or strong staining intensity in ≥ 80% of tumor cells), focal positive (moderate or strong staining intensity in 10 - 79% of tumor cells), or negative (< 10% of tumor cells staining). Histiocytes/macrophages were used as the internal positive control for GPNMB, as they showed high expression of GPNMB.

## Results

### RCCs with high expression of *GPR143* mRNA were enriched for renal neoplasms with *FLCN/TSC/mTOR-TFE* alterations

Previous studies demonstrated that GPR143 is not only highly expressed in melanoma, but also is significantly upregulated in various human carcinomas, including breast cancer, colon cancer and RCC.^18,22^ By interrogating the HPA database, we found *GPR143* mRNA was expressed at high levels in the TCGA-KICH cohort of chRCC (Median: 21.5 protein-coding transcripts per million [pTPM]) and the TCGA-KIRP cohort of pRCC (Median: 12.6 pTPM) (Supplementary Fig. 1), which were ranked as the 2^nd^ and 3^rd^ tumor with high GPR143 expression, respectively, among 31 cohorts of 21 cancer types and just below cutaneous melanoma (Median: 151.4 pTPM).

More interestingly, although the median level of *GPR143* mRNA expression was relatively low (2.3 pTPM) in the TCGA-KIRC cohort of ccRCC, 8 of 12 (66.7%) outlier tumors with high (> 60 pTPM) *GPR143* expression in this TCGA cohort harbored genetic alterations in *FLCN/TSC/mTOR-TFE* pathway (Supplementary file 1). Similarly, 15 of 38 (39.5%) tumors with high (> 60 pTPM) *GPR143* expression in the TCGA-KIRP cohort also contained genetic changes in *FLCN/TSC/mTOR-TFE* pathway (Supplementary file 2). Among these 23 RCCs with high *GPR143* mRNA expression and *FLCN/TSC/mTOR-TFE* pathway aberrations, 13 tumors harbored *TFE3* gene rearrangement with various fusion partners, two tumors harbored *TFEB* gene fusion, and two tumors with *TFEB* amplification, indicating that these 17 tumors should be re-classified as tRCC. The other six tumors with high expression of GPR143 in the TCGA-KIRC and TCGA-KIRP cohorts harbored various mutations in *FLCN/TSC* pathway, including two tumors with *FLCN* deletion, three tumors with *TSC2* mutation and one tumor containing *TSC1* mutation, respectively (Supplementary file 1 and 2).

In contrast, although the average expression level of *GPR143* was higher (30.4 pTPM) in the TCGA-KICH cohort of chRCC when compared to the TCGA-KIRC cohort (12.7 pTPM), no *TFE3/TFEB* amplification, fusion or mutation in *FLCN/TSC1/TSC2* was observed among 6 tumors with the highest (> 60 pTPM) *GPR143* mRNA expression. Only 2 of 26 (7.7%) tumors with relatively high (>30 pTPM) *GPR143* expression displayed *TSC1* mutation (Supplementary file 3). These results indicate many chRCC tumors intrinsically express certain amount of *GPR143* mRNA in the absence of *FLCN/TSC/mTOR-TFE* alterations.

### Renal neoplasms with *FLCN/TSC/mTOR-TFE* alterations frequently expressed high levels of GPR143 protein

To confirm the findings of upregulated expression of GPR143 in renal neoplasms with *FLCN/TSC/mTOR-TFE* alterations, IHC analysis was performed in 23 renal tumors using an antibody against GPR143. Diffuse strong GPR143 immunoreactivity was observed in six of seven (85.7%) tRCCs, while diffuse GPNMB staining was detected in four of seven (57.1%) tRCCs (Fig. 1 and Table 1). One FISH-confirmed *TFE3*-rearranged RCC with abundant clear cytoplasm showed negative for both GPR143 and GPNMB stains (Fig. 1M-1O). It is important to note that although most (five of seven) tRCCs displayed concordant expression pattern between GPR143 and GPNMB, two FISH-confirmed *TFE3*-rearranged RCCs showed diffuse positive for GPR143 and only focal scattered immunoreactivity for GPNMB (Fig. 1G-1L). Interestingly, in the FISH-confirmed *TFE3*-rearranged RCC with biphasic morphology composed of large clear tumor cells and small nested eosinophilic cells clustered in the center, the immunoreactivity for GPNMB was located predominantly in the central pseudorosette structures while the large tumor cells with abundant clear cytoplasm were mainly negative for GPNMB (Fig. 1K). In contrast, both populations of tumor cells were positive for GPR143 labeling (Fig. 1L).

**Figure 1.**
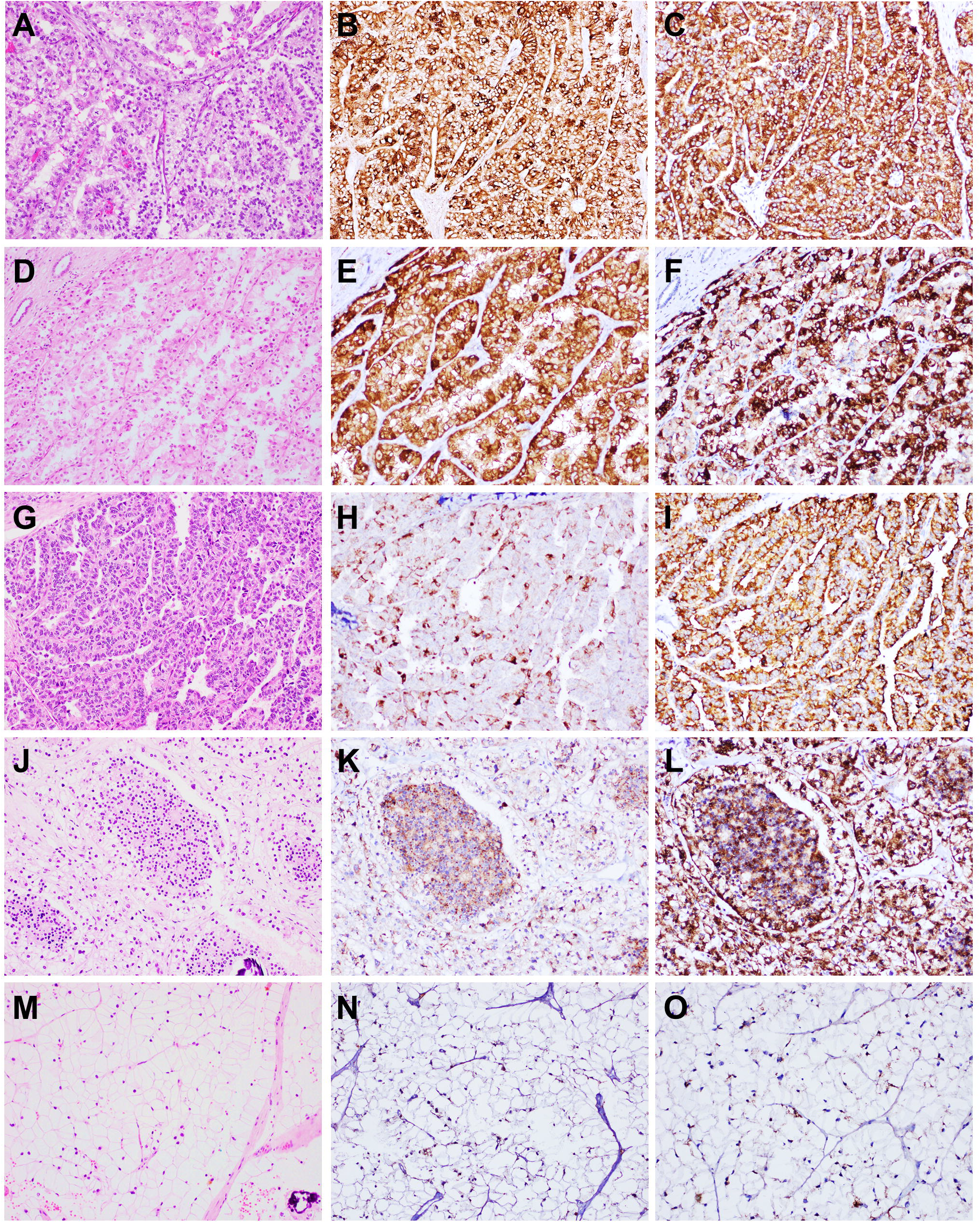
Representative H&E and IHC analysis in MiT/TFE family translocation renal cell carcinomas (tRCCs). (A-C) Diffuse positive GPNMB (B) and GPR143 immunoreactivity (C) in a *TFE3*-rearranged RCC. (D-F) Diffuse positive GPNMB (E) and GPR143 immunoreactivity (F) in a *TFEB*-rearranged RCC. (G-I) Focal GPNMB staining (H) and diffuse GPR143 immunoreactivity (I) in a FISH-confirmed *TFE3*-rearranged RCC. (J-L) Focal GPNMB staining (K) and diffuse GPR143 immunoreactivity (L) in a FISH-confirmed *TFE3*-rearranged RCC with biphasic morphology. (M-O) Negative GPNMB (N) and GPR143 immunoreactivity (O) in a FISH-confirmed *TFE3*-rearranged RCC with abundant clear cytoplasm. Original magnification: 200×.

In four *FLCN*-mutated renal tumors from three BHD syndrome patients with germline *FLCN* mutations, GPR143 and GPNMB immunostains were diffusely positive in three oncocytic tumors, but negative in a ccRCC with a germline *FLCN c*.*1285dup* (p.His429fs) mutation (Fig. 2). Diffuse GPNMB labeling was also found in one of one (100%) ESC with a germline *TSC1* mutation (Fig. 3B), two of two (100%) LOT and one of two (50%) EVT cases (Fig.4), while focal or diffuse GPR143 staining was observed in these five oncocytic tumors (Fig. 3C and Fig. 4). Interestingly, in comparison to strong granular cytoplasmic and cell membrane staining in tRCCs, GPR143 stain in most of these oncocytic tumors exhibited fine granular or dot-like patterns, which may be attributed to its localization in intracellular organelles. In contrast, a *TSC1*-mutated RCC exhibited diffuse strong positive for GPNMB, but negative for GPR143 labeling (Fig. 3D-3F). Furthermore, all six renal AMLs tested were positive for both GPR143 and GPNMB immunostains (Fig. 5A-5I and Table 1). Taken together, these data demonstrated that overlapping expression of GPR143 and GPNMB were observed in most but not all of renal neoplasms with *FLCN/TSC/mTOR-TFE* alterations, and GPR143 immunostain was positive in some cases with negative or focal GPNMB labeling.

**Figure 2.**
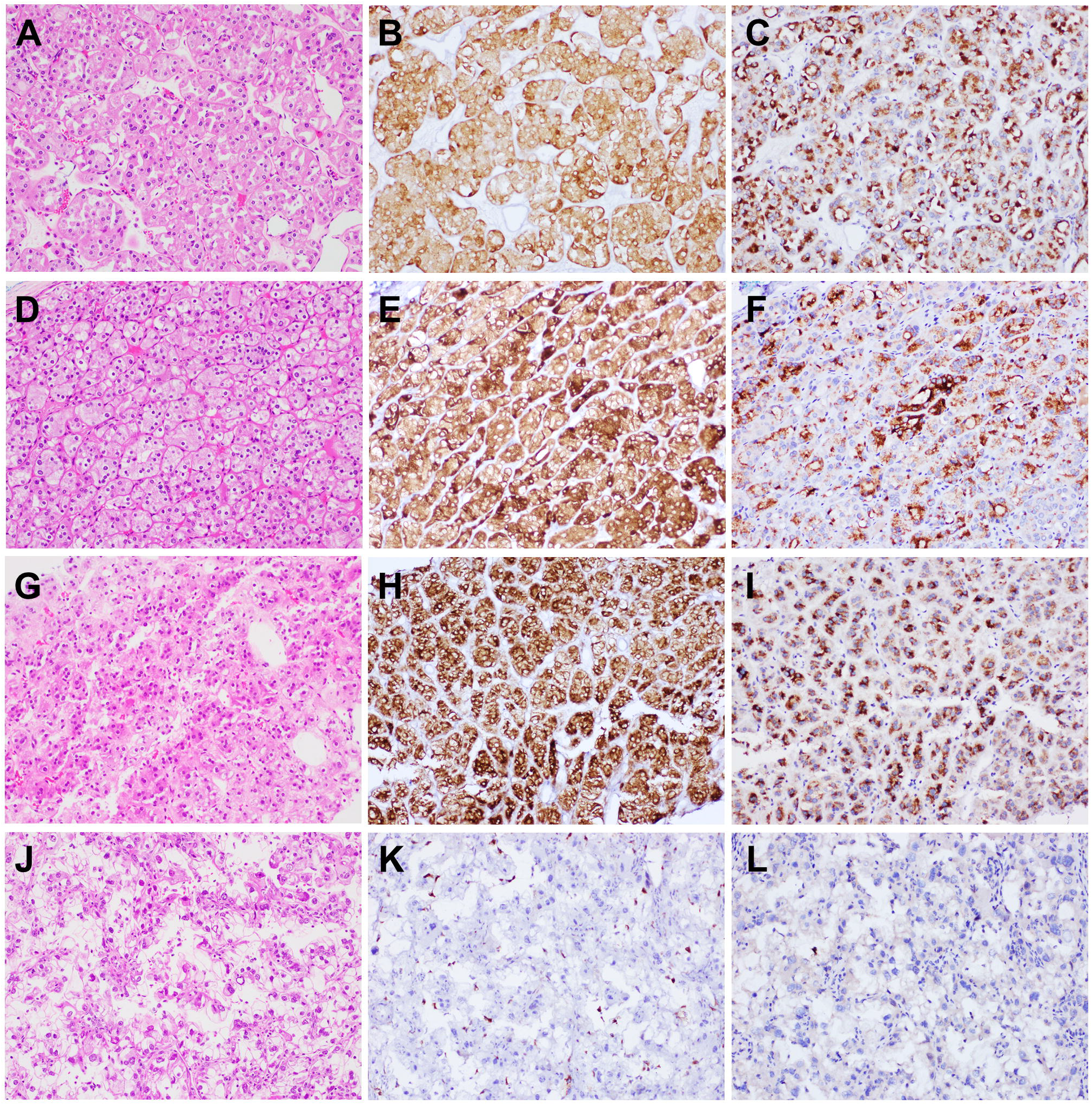
Representative H&E and IHC analysis in *FLCN* mutated renal tumors from three patients with BHD syndrome. (A-F) Diffuse positive GPNMB (B and E) and GPR143 immunoreactivity (C and F) in two HOCTs from one BHD patient with a germline *FLCN* mutation. (G-I) Diffuse positive GPNMB (H) and GPR143 immunoreactivity (I) in an unclassified low grade oncocytic renal neoplasm from one BHD patient with a germline *FLCN* mutation. (J-L) Negative GPNMB (K) and GPR143 immunoreactivity (L) in a ccRCC from one BHD patient with a germline *FLCN* mutation. Original magnification: 200×.

**Figure 3.**
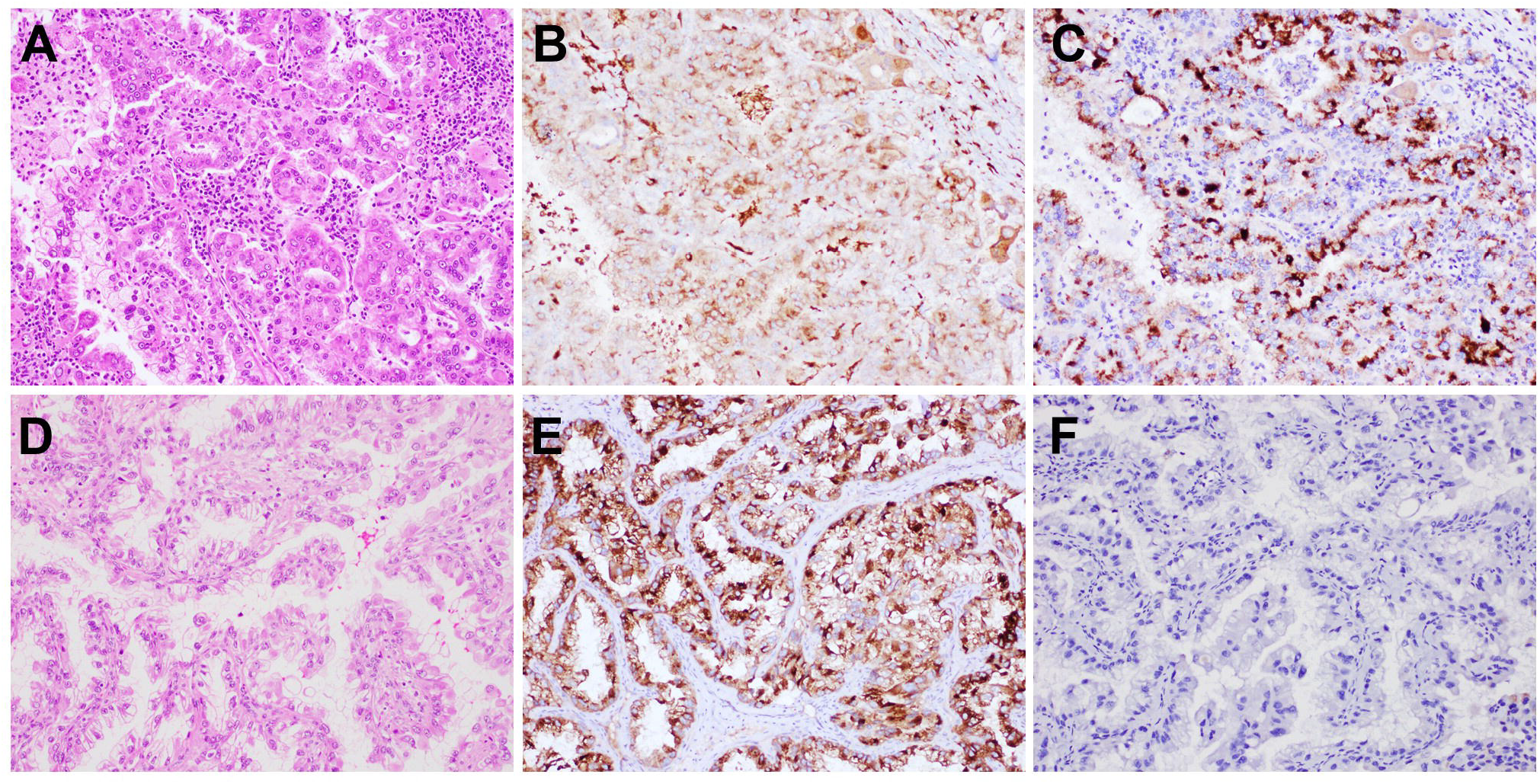
Representative H&E and IHC analysis in *TSC1* mutated renal tumors. (A-C) Diffuse moderate GPNMB staining (B) and diffuse positive GPR143 immunoreactivity (C) in one ESC RCC with a germline *TSC1* mutation. (D-F) Diffuse positive GPNMB staining (E) and negative GPR143 immunoreactivity (F) in an unclassified *TSC1* mutated RCC from a patient with TSC and a germline *TSC1* mutation. Original magnification: 200×.

**Figure 4.**
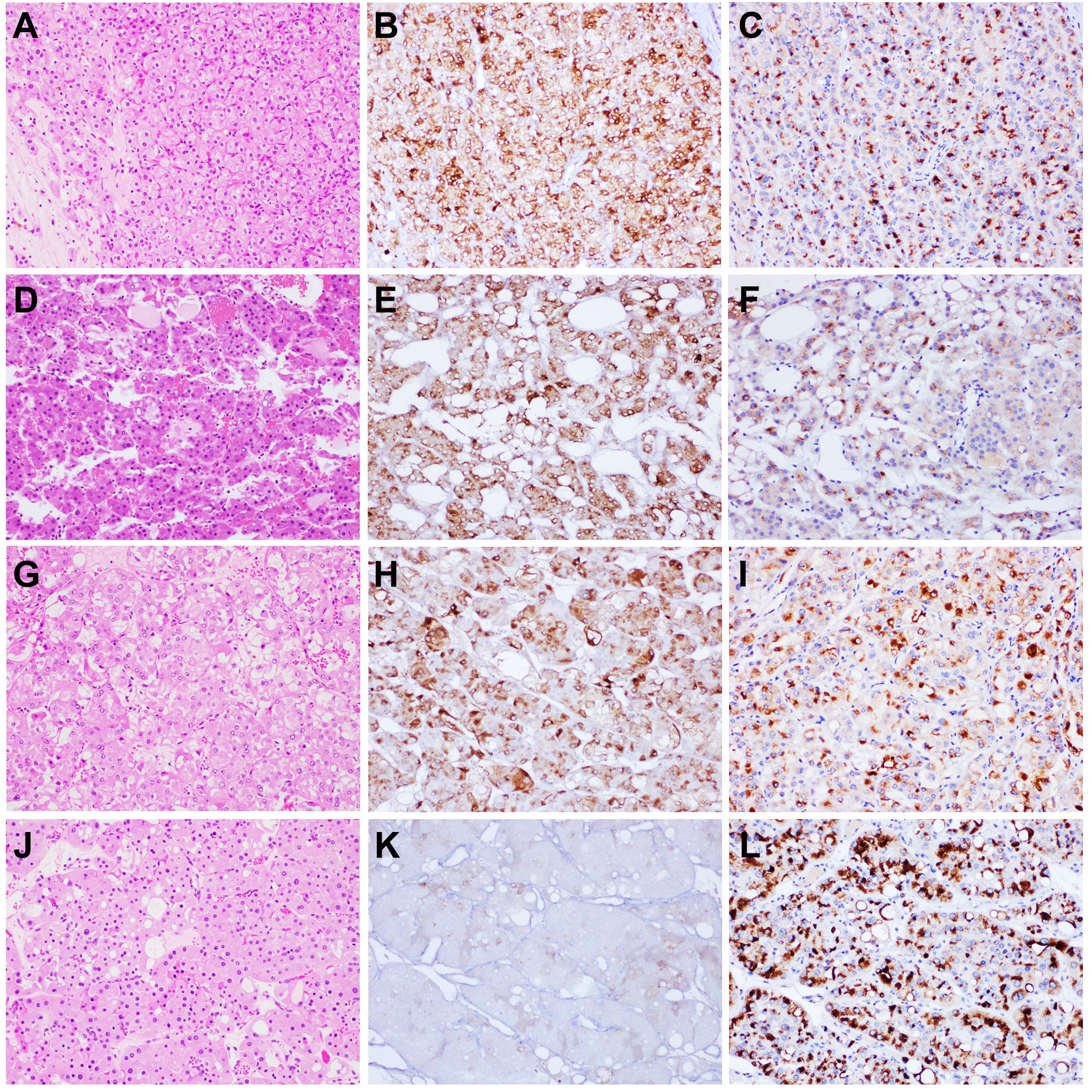
Representative H&E and IHC analysis in oncocytic renal neoplasms. (A-F) Diffuse positive GPNMB (B and E) and focal GPR143 immunoreactivity (C and F) in two cases of LOT. (G-I) Diffuse positive GPNMB (H) and GPR143 immunoreactivity (I) in one EVT case. (J-L) Negative GPNMB (K) and diffuse positive GPR143 immunoreactivity (L) in one EVT case. Original magnification: 200×.

**Figure 5.**
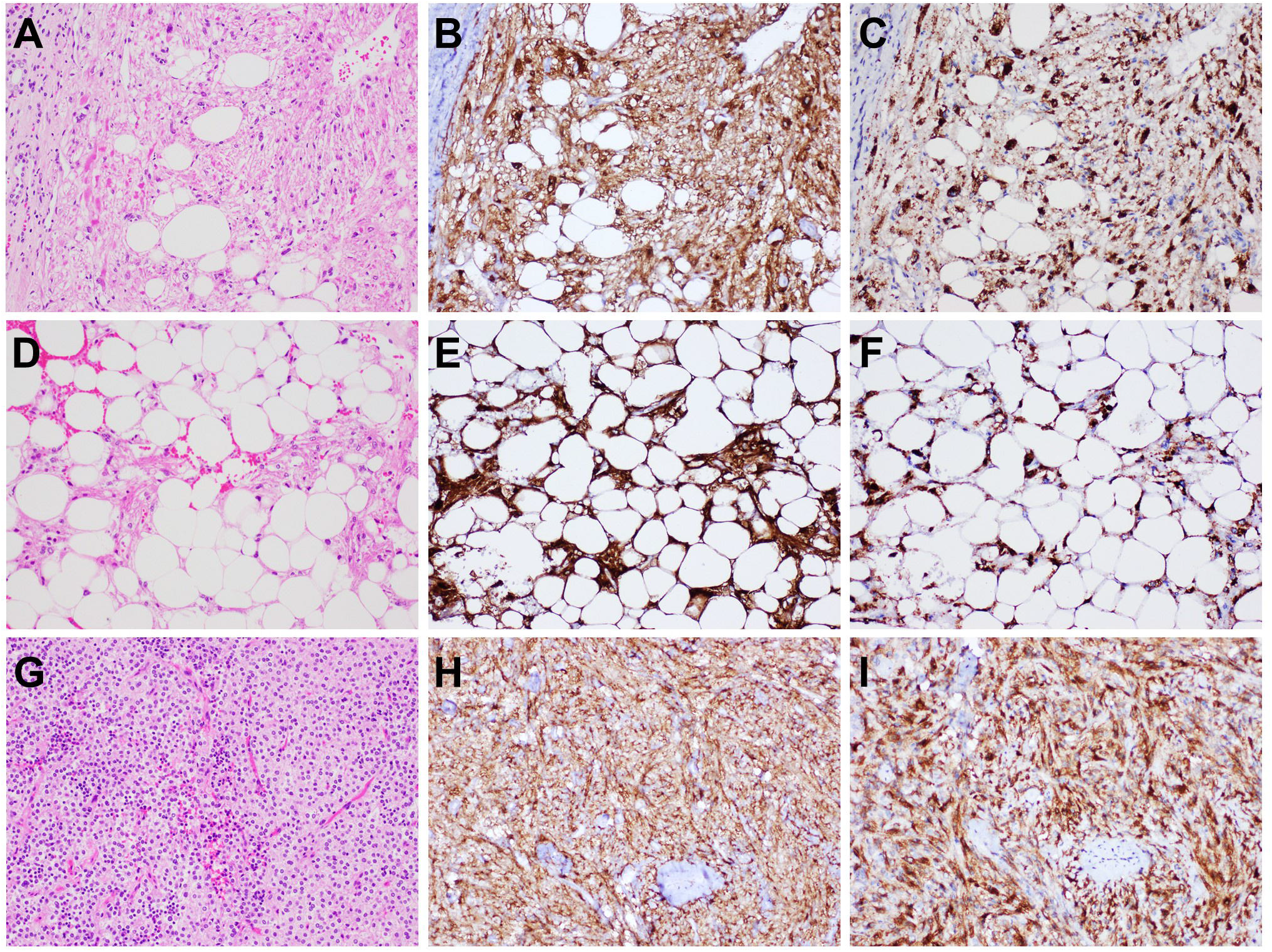
Representative H&E and IHC analysis in renal angiomyolipomas (AMLs). (A-C) Diffuse positive GPNMB (B) and GPR143 immunoreactivity (C) in a renal AML from a patient with TSC. (D-F) Positive GPNMB (E) and GPR143 immunoreactivity (F) in a lipid-rich AML. (G-I) Positive GPNMB (H) and GPR143 immunoreactivity (I) in a renal epithelioid AML. Original magnification: 200×.

To evaluate the specificity of GPR143 stain for renal neoplasms with *FLCN/TSC/mTOR-TFE* alterations, GPR143 IHC analysis was performed in a collection of three most common histological types of RCC. Only 1 of 17 (5.9%) ccRCC showed focal moderate GPR143 stain in < 30% tumor area. However, one of seven (14.3%) pRCC cases exhibited diffuse strong immunoreactivity for both GPR143 and GPNMB, and negative for TFE3, HMB45 and MART-1 stains (Supplementary Fig. 2 and data not shown). Consistent with the upregulated expression of *GPR143* mRNA in the TCGA-KICH cohort, two of eight (25%) chRCC tested displayed diffuse positive for GPR143, while five of eight (62.5%) chRCC showed diffuse GPNMB immunoreactivity (Supplementary Fig. 3). These results suggest neither GPR143 nor GPNMB are specifically expressed in tRCCs or *FLCN/TSC* mutated renal neoplasms, and a significant fraction of chRCC could display positivity for these two biomarkers.

## Discussion

tRCC is a group of molecularly defined RCCs, consisting of *TFE3*-rearranged RCC, *TFEB*-rearranged RCC and *TFEB*-amplified RCC. The diagnosis of tRCCs mainly relies on FISH analyses, which are expensive and not easily available in most clinical laboratories. It has been shown that IHCs using several biomarkers, including TFE3, cathepsin K and GPNMB, are useful ancillary screening tests to identify tRCCs. However, the sensitivities of these biomarkers are not very high and the interpretation of ambiguous TFE3 and GPNMB immunostains could be challenging.^1,2,5,23^ It has been proposed that only diffuse GPNMB immunoreactivity is predictive of *FLCN/TSC/mTOR-TFE* alterations in renal tumors.^1,2,5^ In this study, we demonstrated that a large fraction of tumors with high levels of *GPR143* expression in TCGA-KIRC and TCGA-KIRP cohorts harbored *TFE3/TFEB* alterations. Consistently, significant overexpression of *GPR143* in tRCC was found by RNA-seq analysis of 15 *TFE3*-rearranged tRCC samples as well as TFE3 ChIP-seq test using the UOK146 cell line harboring a *PRCC-TFE3* gene fusion.^24^ To confirm upregulated GPR143 protein in tRCC, we performed IHC analysis with an antibody against GPR143 and demonstrated positive GPR143 immunoreactivity in the majority of tRCC cases tested. Moreover, two FISH-confirmed *TFE3*-rearranged RCCs with focal scattered immunoreactivity for GPNMB showed diffuse positive for GPR143. Our results suggest that complementary to the aforementioned existing markers, GPR143 could serve as a useful adjunct marker to increase the sensitivity for tRCC screen and diagnosis. Multiplex IHC analysis using a cocktail of antibodies against these biomarkers could be an ideal diagnostic tool to efficiently screen tRCCs with a lower false negative rate and simultaneously save the limited biopsy tissues for subsequent molecular tests.

In addition, we showed that similar to GPNMB IHC analysis, GPR143 immunoreactivity was observed in the majority of *FLCN* mutated renal tumors and renal neoplasms with high frequencies of *TSC/mTOR* signal pathway aberrations, indicating that GPR143 could also function as a surrogate marker for *FLCN/TSC/mTOR* alterations in renal tumors. However, we also found that a significant fraction of chRCC cases displayed diffuse positive labeling for these two biomarkers. Although aberrant TSC/mTOR signaling pathway has been implicated in the pathogenesis and disease progression in a subset of chRCC,^25,26^ additional mechanisms independent of *FLCN/TSC* mutations or mTOR pathway activation, such as loss of transcriptional repressors or epigenetic changes, could play roles in upregulation of GPR143 and GPNMB in some chRCC and other oncocytic renal tumors. Moreover, it is also possible that the granular and dot-like staining pattern of GPR143 observed in most of *FLCN/TSC/mTOR*-related oncocytic renal tumors could be partially attributed to increased mitochondria or dysfunctional lysosome and other intracellular organelles. Therefore, further studies with larger cohorts of chRCC and other oncocytic renal neoplasms with *FLCN/TSC/mTOR* alterations are warranted to evaluate the specificity of these biomarkers for *FLCN/TSC/mTOR* alteration prediction.

In this study, we showed that both GPNMB and GPR143 immunostains yielded negative results in one FISH-confirmed *TFE3*-rearranged RCC with abundant clear cytoplasm. These two biomarkers were also negative in one ccRCC with a germline *FLCN* frameshift mutation. Consistently, a recent study described positive GPNMB immunoreactivity in only 25% (two of eight) of ccRCC cases with coexisting *VHL* and *TSC*/*mTOR* mutations.^9^ Taken together, these results indicate that relative to the high predictive value in oncocytic renal neoplasms, both GPNMB and GPR143 immunoreactivity may be insensitive indicators for *FLCN/TSC/mTOR*-*TFE* aberrations in renal tumors with clear cytoplasm. The underlying mechanisms for lower positive rates of GPNMB and GPR143 stains in these tRCCs or mTOR-activated renal tumors with clear cell morphology remain unclear, although accumulation of glycogen and lipids, altered lysosomal/organelle trafficking or increased mitochondrial autophagy could play contributary roles.

*TFEB*-altered RCCs are much less common than *TFE3*-rearranged RCCs. Among *TFEB*-altered RCCs, the incidence of *TFEB-*amplified RCC seems to be relatively lower compared to that of *TFEB*-rearranged RCC. Currently, approximately 100 cases of *TFEB-*amplified RCC have been reported in the literature.^27-29^ Due to the small cohort of our study and the rare incidence of *TFEB-*altered RCCs, we only included a *TFEB*-rearranged RCC case, which exhibited diffuse GPNMB and GPR143 labeling. Although there was no *TFEB-*amplified RCC in this study, it is very likely that *TFEB-*amplified RCC cells will display positive GPR143 immunoreactivity because two *TFEB-*amplified RCC cases in TCGA cohorts expressed high levels of *GPR143* mRNA (Supplementary file 1 and 2). It has been shown in a recent study that negative or patchy/equivocal GPNMB immunoreactivity was observed in 36.4% (4 of 11) of *TFEB-*amplified RCCs.^2^ Therefore, further study is needed to compare the positive rates of GPNMB and GPR143 immunostains in this specific subtype of *TFEB*-altered RCCs.

In addition to the low number of tRCC cases and *FLCN/TSC/mTOR*-related renal tumors, the other limitations of our study include the retrospective nature of study from a single tertiary care center and the lack of molecular confirmation of *TSC/mTOR-TFE* aberrations in a significant proportion of this cohort. However, given that GPR143 immunostain was positive in several renal tumors with negative or focal GPNMB staining, we believe that our findings of GPR143 expression in renal neoplasms with *FLCN/TSC/mTOR-TFE* alterations should be of clinical significance. These results not only provide a novel adjunct marker for precise diagnosis of renal tumors with overlapping morphology, but also have potential values for subclassification and management of kidney cancer patients based on the aberrantly activated signal pathways. Due to the high frequencies of positive GPR143 immunoreactivity in *TFE3/TFEB*-rearranged RCC and renal AML, it will be interesting to study GPR143 expression in other *TFE*-altered conditions, such as alveolar soft part sarcomas and non-renal perivascular epithelioid cell tumor (PEComa). Since GPR143 is a MiT/TFE family of transcriptional target implicated in melanosome biogenesis and maturation,^14-16^ the potential clinical application of GPR143 immunostain could be extended to several cutaneous and soft tissue disorders upon optimization and validation, which could provide another useful melanocytic differentiation marker to facilitate the diagnosis of various melanocytic lesions and clear cell sarcoma of soft tissue.

In summary, by interrogating the HPA and cBioPortal databases, high frequencies of *FLCN/TSC/mTOR-TFE* alterations were identified among RCCs with high levels of GPR143 expression in two TCGA kidney cancer cohorts. Similar to GPNMB labeling, GPR143 immunostain was positive in the majority of tRCC cases and renal tumors with *FLCN/TSC/mTOR* alterations, suggesting that GPR143 could function as another surrogate marker for *FLCN*/*TSC/mTOR-TFE* signal pathway aberrations in certain renal tumors. Despite the overlapping expression of GPR143 and GPNMB in most renal neoplasms with *FLCN/TSC/mTOR-TFE* alterations, diffuse GPR143 immunoreactivity was found in some cases with negative or focal GPNMB staining. These results indicate GPR143 could serve as a useful adjunct marker to improve the sensitivity for screening renal tumors with *FLCN/TSC/mTOR-TFE* alterations.

## Data Availability

All data produced in the present study are available upon reasonable request to the authors.

## Figure legends

**Supplementary figure 1.**
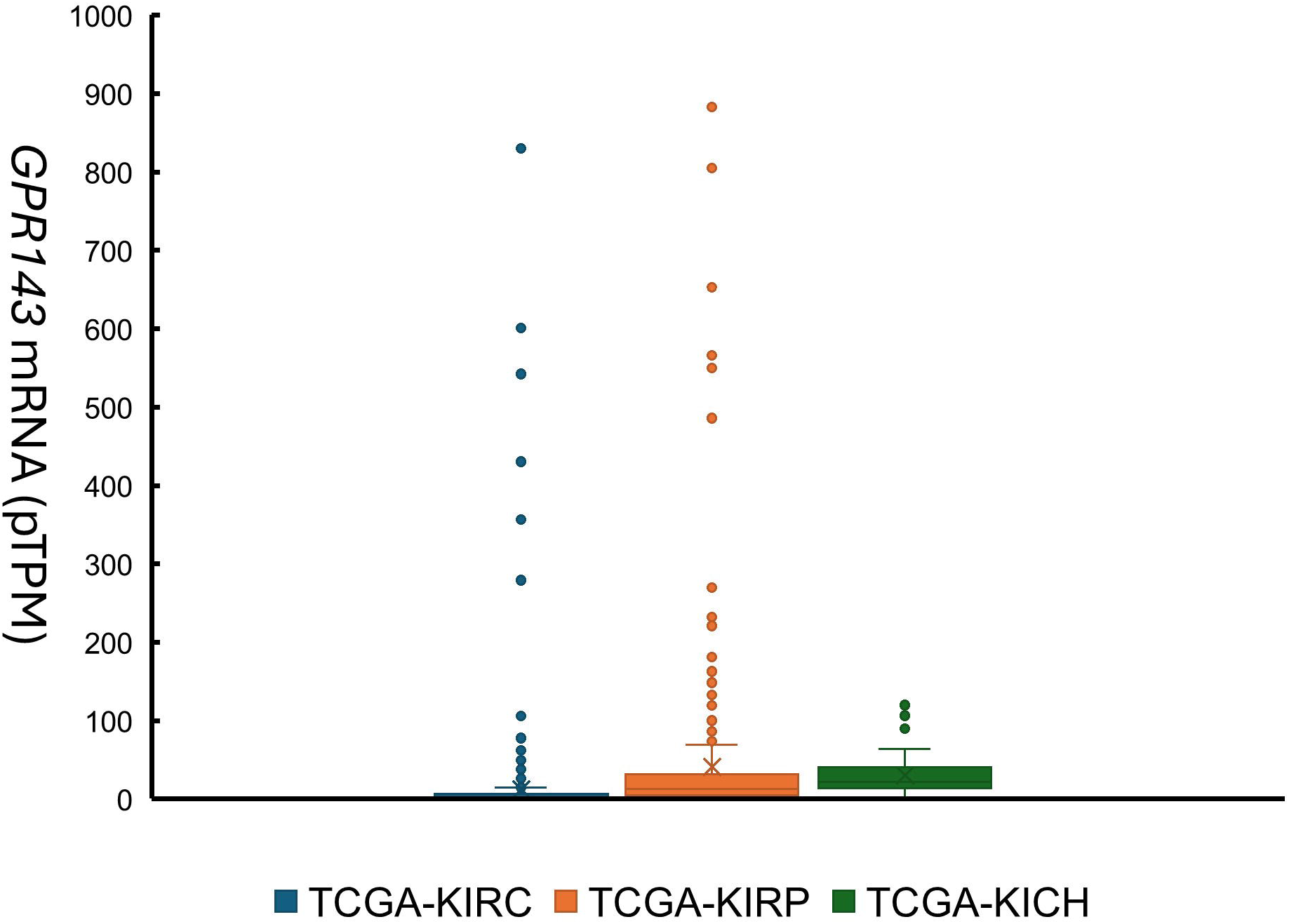
Expression of *GPR143* RNA in renal cell carcinomas in three TCGA kidney cancer cohorts. TCGA-KIRC: Clear cell RCC; TCGA-KIRP: Papillary RCC; TCGA-KICH: Chromophobe RCC.

**Supplementary figure 2.**
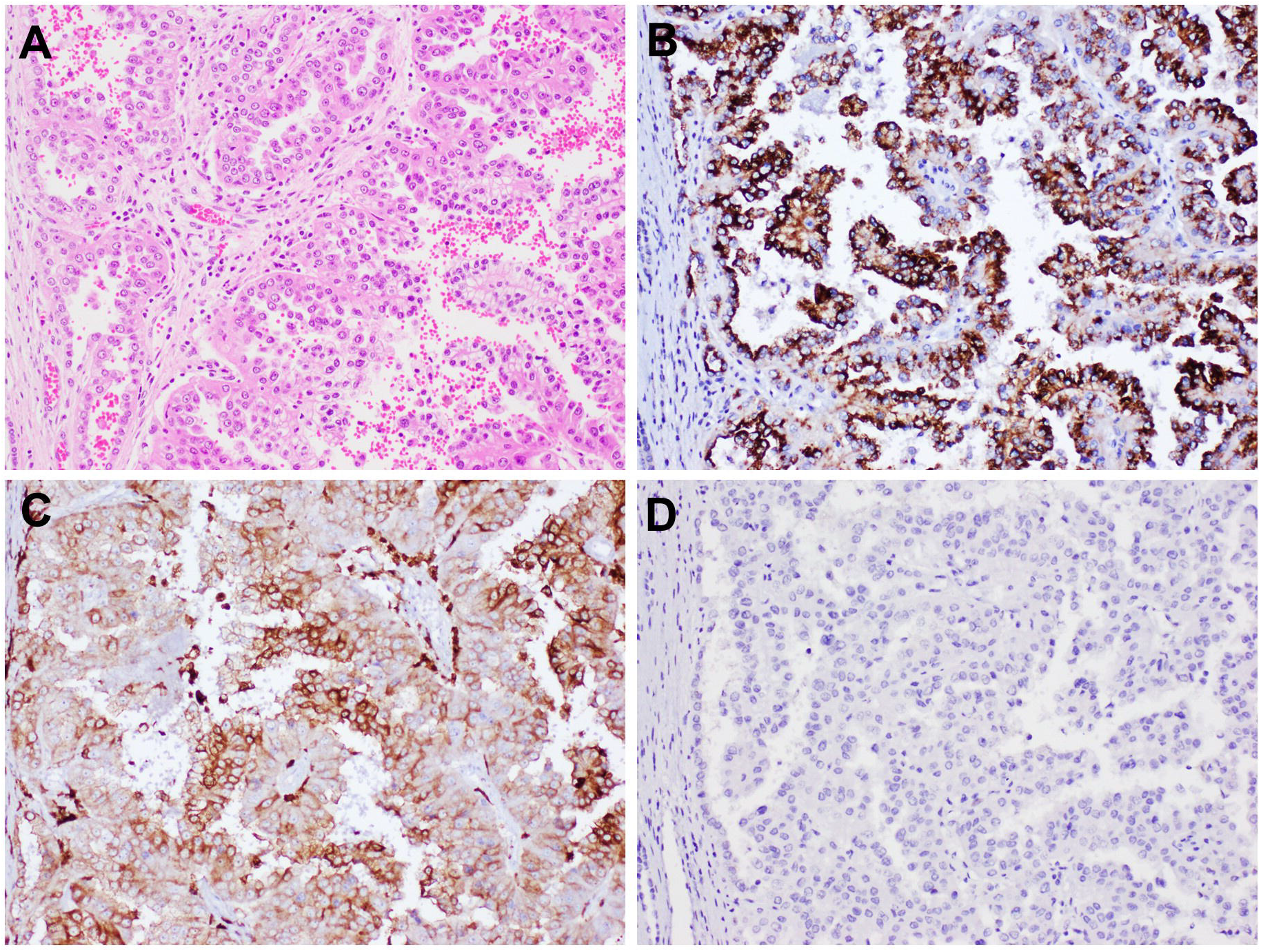
Representative H&E and IHC analysis in a papillary renal cell carcinoma. (A) H&E stain. (B) Diffuse positive GPR143 staining. (C) Diffuse positive GPNMB immunoreactivity. (D) Negative TFE3 staining. Original magnification: 200×.

**Supplementary figure 3.**
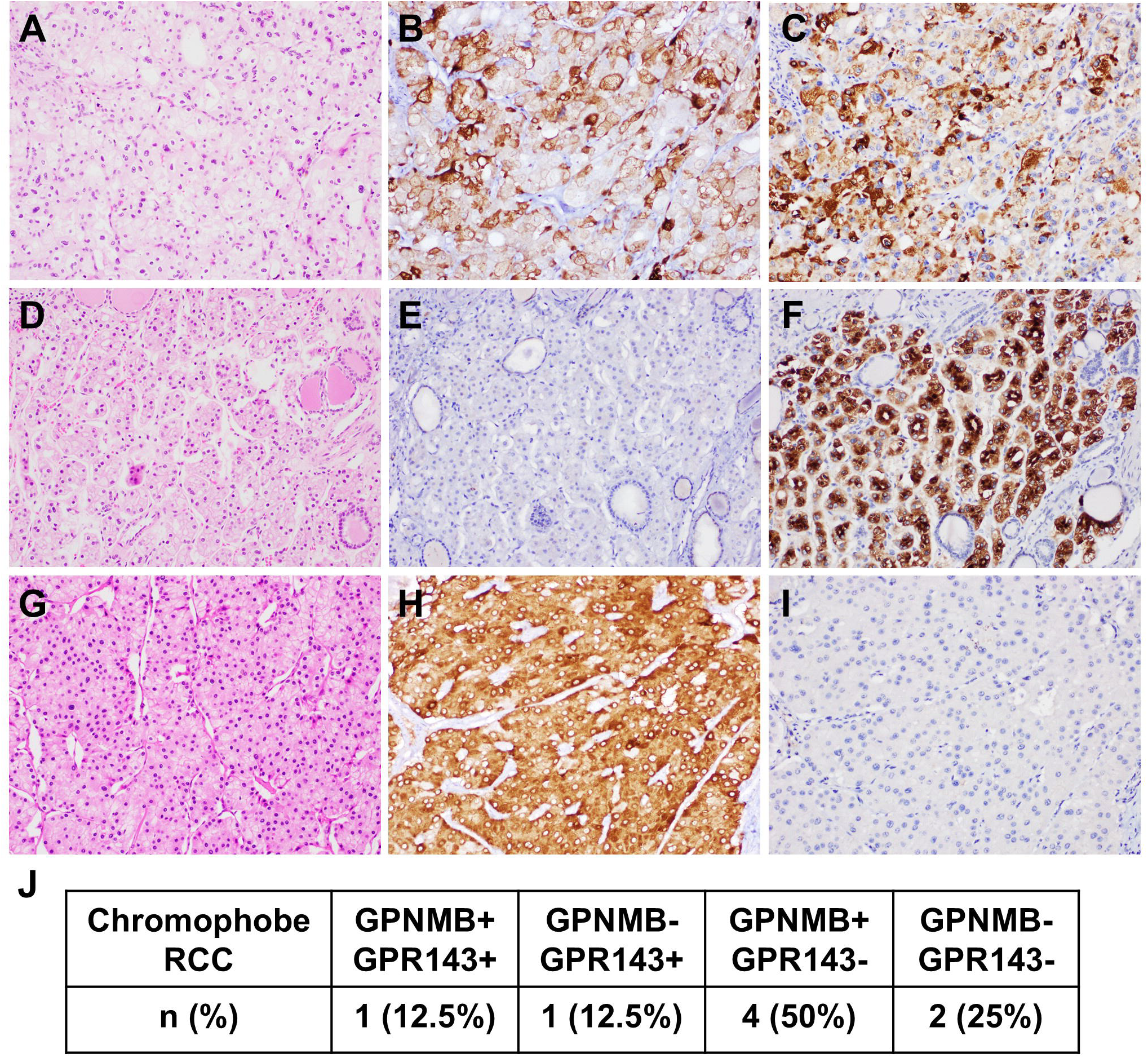
Representative H&E and IHC analysis in chromophobe renal cell carcinomas (chRCCs). (A-C) Diffuse positive GPNMB (B) and GPR143 immunoreactivity (C) in one chRCC. (D-F) Negative GPNMB staining (E) and positive GPR143 immunoreactivity (F) in a chRCC. (G-I) Diffuse positive GPNMB immunoreactivity (H) and negative GPR143 staining (I) in a chRCC. Original magnification: 200×. (J) Summary of GPNMB and GPR143 immunostaining patterns in eight chRCCs.

